# Evolving cartilage strain with pain progression and gait: a longitudinal study post-ACL reconstruction at six and twelve months

**DOI:** 10.1101/2024.09.08.24313289

**Authors:** Emily Y. Miller, Timothy Lowe, Hongtian Zhu, Woowon Lee, Pablo F. Argote, Danielle Dresdner, James Kelly, Rachel M. Frank, Eric McCarty, Jonathan Bravman, Daniel Stokes, Nancy C. Emery, Corey P. Neu

## Abstract

**Background:** Anterior cruciate ligament (ACL) injuries are prevalent musculoskeletal conditions often resulting in long-term degenerative outcomes such as osteoarthritis (OA). Despite surgical advances in ACL reconstruction, a significant number of patients develop OA within ten years post-surgery, providing a patient population that may present early markers of cartilage degeneration detectable using noninvasive imaging.

**Purpose:** This study aims to investigate the temporal evolution of cartilage strain and relaxometry post-ACL reconstruction using displacement under applied loading MRI and quantitative MRI. Specifically, we examined the correlations between MRI metrics and pain, as well as knee loading patterns during gait, to identify early candidate markers of cartilage degeneration.

**Materials and Methods:** Twenty-five participants (female/male = 15/10; average age = 25.6 yrs) undergoing ACL reconstruction were enrolled in a prospective longitudinal cohort study between 2022 and 2023. MRI scans were conducted at 6- and 12-months post-surgery, assessing T2, T2*, and T1ρ relaxometry values, and intratissue cartilage strain. Changes in pain were evaluated using standard outcome scores, and gait analysis assessed the knee adduction moment (KAM). Regressions were performed to evaluate relationships between MRI metrics in cartilage contact regions, patient-reported pain, and knee loading metrics.

**Results:** Increases in axial and transverse strains in the tibial cartilage were significantly correlated with increased pain, while decreases in shear strain were associated with increased pain. Changes in strain metrics were also significantly related to KAM at12 months.

**Conclusions:** Changes in cartilage strain and relaxometry are related to heightened pain and altered knee loading patterns, indicating potential early markers of osteoarthritis progression. These findings underscore the importance of using advanced MRI for early monitoring in ACL-reconstructed patients to optimize treatment outcomes, while also highlighting KAM as a modifiable intervention through gait retraining that may positively impact the evolution of cartilage health and patient pain.

**Key Results:**

1. Increased axial and transverse strains in the tibial cartilage from 6 to 12 months post-ACL reconstruction were significantly correlated with increased pain, suggesting evolving changes in cartilage biomechanical properties over time.
2. Decreases in shear strain in inner femoral and central tibial cartilage regions were linked to increased pain, indicating alterations in joint loading patterns.
3. Decreases in shear strain in the inner femoral cartilage were significantly associated with decreased 12-month knee adduction moment (KAM), a surrogate for medial cartilage knee loading during walking.

## Introduction

Anterior cruciate ligament (ACL) injuries are a common orthopedic condition that can have long-term consequences for patients. Despite advances in ACL reconstructive surgical techniques, a significant number of patients develop osteoarthritis (OA) within ten years of ACL injury (1). Osteoarthritis is a degenerative joint disease characterized by cartilage degradation and structural changes impacting the entire knee joint. As this condition progresses, patients frequently experience knee pain, stiffness, and a lower quality of life, often necessitating total knee arthroplasty at later stages. Alterations in knee loading patterns are theorized to intensify joint degeneration as osteoarthritis develops (2–5). These biomechanical changes can trigger breakdown of the articular cartilage, as well as structural modifications like bone marrow edema, synovitis, and osteophyte formation within the joint. It is crucial to identify patients at high risk of future osteoarthritis early in the postoperative period, to allow intervention before irreparable damage occurs. However, our understanding of the underlying mechanisms that initiate cartilage degeneration following ACL injury and reconstruction, and how those differ on a personalized basis, is still incomplete.

Magnetic resonance imaging (MRI) has emerged as a promising modality that can provide insights beyond the conventional radiographic methods used to diagnose OA. Although existing semi-quantitative scoring systems provide morphological information about cartilage, they are unable to detect the subtle matrix alterations and biomechanical property changes that characterize early osteoarthritic cartilage (6–8). Recent research has employed quantitative MRI (qMRI) techniques and displacements under applied loading MRI (dualMRI) in healthy and ACL-reconstructed individuals to quantify intratissue strain and relaxometry values (9–12). Building upon this foundation, our study investigates the temporal evolution of cartilage strain and relaxometry in a clinical cohort undergoing ACL-reconstruction (ACLR). Through longitudinal assessment of dualMRI-derived strain and qMRI-derived relaxometry from 6 to 12 months post-ACLR, and correlation with patient-reported outcomes and measures of knee loading through gait analysis, we aim to identify early markers of cartilage degeneration and their clinical implications as well as offer valuable insights into the progressive nature of cartilage remodeling. The study had three objectives. First, we studied the relationship between medial articular cartilage dualMRI-derived strains and qMRI relaxometry at 6- and 12-month post-ACLR. Second, we investigated how changes in participant pain are related to changes in MRI over this period. Third, in a subset of participants, we examined how strain changes functionally relate to human motion, using peak knee adduction moment (KAM) at 12-months as a surrogate for knee loading.

## Materials and Methods

### Participants

Between 2022 and 2023, participants were enrolled in a longitudinal cohort study at the University of Colorado Anschutz orthopedic clinic following unilateral ACL reconstructive surgery. The University of Colorado IRB approved the study, and all participants gave informed consent. Inclusion criteria included individuals aged 18 to 40 who had ACL surgery within three months of injury, with no prior symptomatic knee pathologies. Exclusion criteria included previous ACL surgery on either limb or cartilage chondral defect surgery. Participants who sustained further knee injuries between six- and twelve-months post-surgery were also excluded. The ACL reconstruction used either a bone-patella tendon-bone autograft or a quadriceps tendon autograft. Rehabilitation adherence was not tracked, but all followed the MOON ACL protocol (13). MRI scans were conducted at approximately six- and twelve-months post-surgery, before which they completed the Knee Injury and Osteoarthritis Outcome Score (KOOS) and Western Ontario and McMaster Universities Arthritis Index (WOMAC) questionnaires. Pain percent change was calculated by scaling and averaging the KOOS and WOMAC pain subscores from six to twelve months.

### Loading Protocol of Human Knee

The study utilized a previously validated MRI-compatible loading apparatus to apply a compressive load to the tibiofemoral cartilage during the DENSE sequence (12). This apparatus directly applied a load perpendicular to the ankle joint, inducing a varus moment at the knee, and causing the compressive force at the medial cartilage condyle equivalent to 0.5 times the participant’s body weight. Due to the nature of the DENSE sequence, the loading was applied useing a cyclic regime mimicking a walking cadence, with 1-second load and unload intervals (9,11,12).

### MRI Scanning Protocol and Processing

The knee imaging protocol remained consistent throughout all scanning sessions. Initially, a fast gradient echo MR image sequence was utilized for localization, which was followed by a 3D double echo steady state (DESS) acquisition, quantitative T2, T2*, and T1ρ measurements, and a displacement encoding with stimulated echoes (DENSE) sequence during cyclic varus load of the knee joint. Additionally, to reduce the viscoelastic effect of cartilage, participants underwent an eight-minute cyclic preconditioning load before DENSE acquisition (10,11,13). The imaging procedures were carried out using a Siemens 3T Prisma MRI system equipped with a 15-channel quadrature knee coil. Segmentation of the cartilage into femoral and tibial regions of interest (ROIs) was achieved using a semi-automatic algorithm. Participants were excluded at this stage if motion or metal artifact obscured the medial cartilage ROI. Strain calculations, including Green-Lagrange axial, transverse, and shear strains, were determined from smoothed displacements using the deformation gradient tensor (13–17) within each ROI. For the six-month data set, cartilage ROIs were warped through standard nonrigid image registration using anatomical knee landmarks to align with those at twelve months for each participant (18). Subsequently, a pixel-wise percent change was calculated for each MRI metric and for each participant. Sub-regions were established within the ROIs based on cartilage contact, with identification and segmentation achieved by fitting a spline to the middle of the region of interest and then calculating the length of the spline from the most medial to the most lateral extents. Each spline was then divided into thirds in the medial to lateral direction to define the extent of each region, resulting in the identification of the most medial cartilage-cartilage contact region, a central cartilage-cartilage contact region, and a more lateral cartilage-meniscus contact area (10,11). To conduct statistical analyses, raw and percent change averages for each MRI metric were computed within each cartilage subregion.

### Markerless motion capture and musculoskeletal simulation

During their twelve-month visit, a subset of the participants were selected to undergo gait analysis. All 9 participants performed 3 over-ground walking trials at a self-selected pace. immediately prior to their MRI scan using OpenCap, a markerless motion capture system(19). OpenCap was sampled at 60Hz using 3 commercial iPads (iPad Pro, Apple Inc., Cupertino, CA, USA). The iPads were positioned 1.5m off the ground, 3m from the center of the capture area, and at ±45° and 0^0^, where 0° faces the participant. A precision-manufactured, 720 x 540 mm checkerboard was used for computing the extrinsic parameters during OpenCap’s camera calibration step, as per the operating procedures provided by OpenCap. After recording trials, data was uploaded to the cloud. Trial data then went through automated extraction, pose estimation, time synchronization, and 3D anatomical marker set derivation. 3D kinematics were then computed from marker trajectories using invers kinematics and a musculoskeletal model with biomechanical constraints(19–21). Finally, OpenCap estimated dynamics using muscle-driven tracking simulations of joint kinematics. Peak KAM was then evaluated during the loading phase. Technical details of OpenCap’s software and processing can be found at www.opencap.ai and reported in Uhlrich et al. “OpenCap: human movment dynamics from smartphone videos” (19). T-tests were used to determine that there were no differences in age, BMI, or patient-reported outcome pain scores between the subgroup of nine and the greater group of 25.

### Statistical Analysis

Statistical analyses were carried out using SAS Version 9.4 (SAS Institute) with a 2-sided significance level of ɑ=0.05 by author E.Y.M. Any data that did not meet the assumption of normality as determined by the Shapiro-Wilk test underwent logarithmic transformation using standard methods. Following this, the relationships between 6-month and 12-month MRI data within each cartilage subregion were assessed using simple linear regression (22). Additionally, the associations between changes in pain and MRI values for each cartilage subregion were evaluated using general linear models. A separate model was developed for each MRI metric and subregion, with sex considered as a fixed categorical effect and BMI as a covariate. The residuals from each model were assessed to test for normality and homoskedasticity (23). General linear models were also employed to investigate the correlation between peak knee adduction moment and strain change in the subgroup that underwent gait analysis (23).

## Results

### Participant characteristics

In total, 27 participants took part in both the six-month and twelve-month scans. Two individuals were excluded from the study pool due to motion or metal artifact interference with the cartilage region of interest (Figure 1). Thus, the final sample size comprised 25 participants, including 10 males and 15 females. For the six-month scan, the average participant age was 25.6±5.8 years, with a mean body mass index (BMI) of 24.2±4.1. The demographic and surgical characteristics of the participants are detailed in Table 1. Notably, the subgroup chosen for OpenCap gait analysis encompassed 9 participants, consisting of 6 females and 3 males. Their average age was 23.0±3.1, with an average BMI of 26.33±6.1.

**Figure 1:**
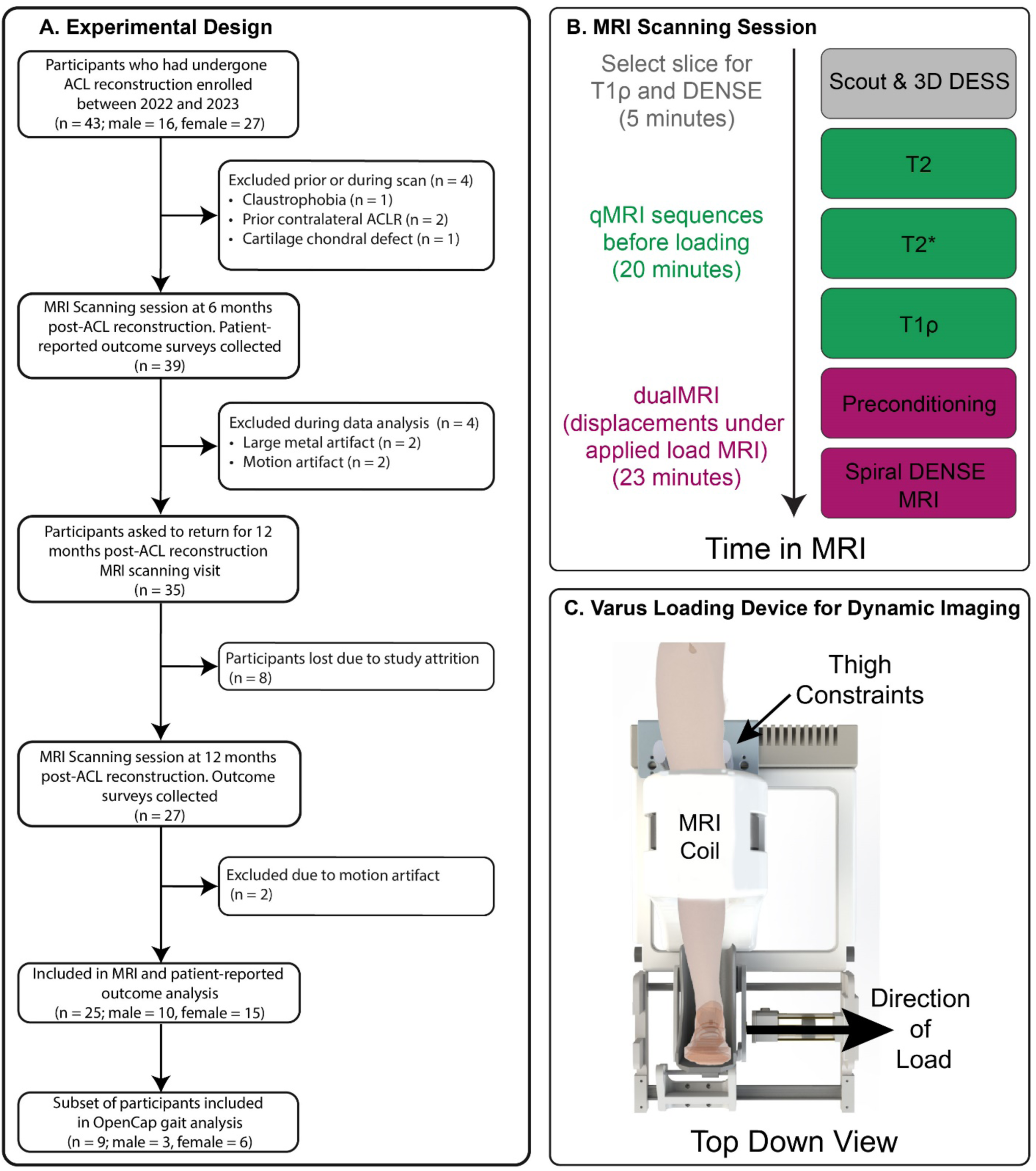
Flow diagram of participant selection and inclusion criteria. The experimental design included recruiting in the orthopedic clinic, 43 participants who had received a unilateral ACL reconstruction. Four participants were excluded prior to the first MRI scan at six months post-ACL reconstruction due to claustrophobia, prior ACLR on the contralateral limb, or cartilage chondral defect. After scanning, four more participants were excluded from analysis due to the presence of large metal artifact in the cartilage region of interest, or motion artifact that obscured analysis. All participants who participated in the scanning session at six-months post ACL reconstruction were invited back to participant in a second scan at twelve-months post ACL reconstruction. 27 participants elected to participate in this scanning session. Two participants were then excluded due to motion artifact. A subset of the 25 participants was selected to participate in gait analysis. B. The MRI scanning session was the same at both the six-month and twelve-month visits. The scanning session included a scout and 3D (double echo steady state (DESS) sequences to visualize knee anatomy and chose a slice that maximizes tibiofemoral contact for both DENSE and T1ρ acquisition. Relaxometry measurements (T2, T2*, T1ρ) were then acquired. C. To apply a cyclic load to the medial knee cartilage during DENSE acquisition, an MRI-compatible varus loading apparatus was used. To minimize the effect of cartilage viscoelasticity, 8 minutes of load were applied in a preconditioning period before DENSE acquisition. ACL, anterior cruciate ligament. MRI, magnetic resonance imaging.

**Table I:**
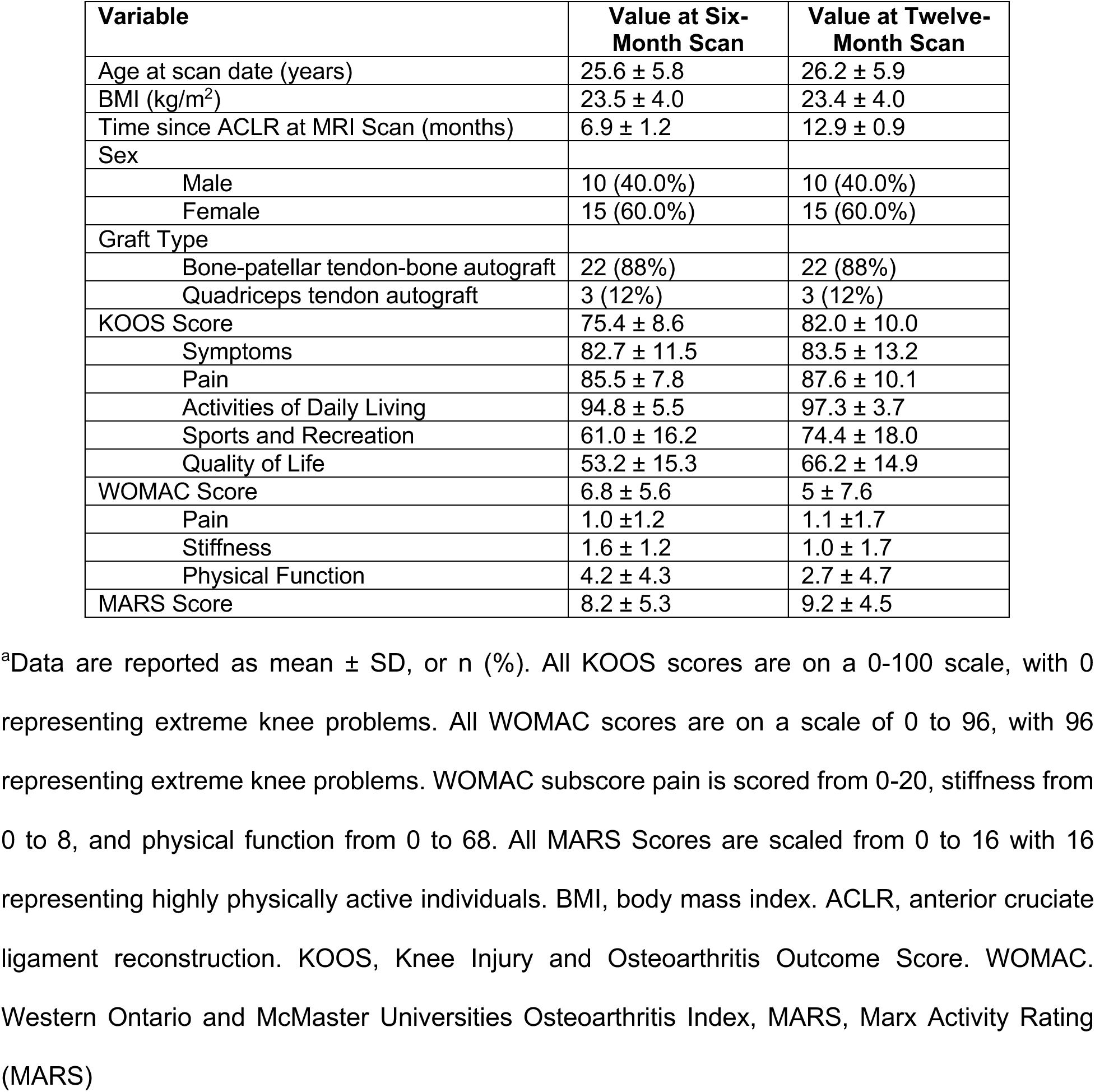
Participant demographic, surgical characteristics, and patient-reported outcomes within 25 subjects included for all analyses.

| Variable | Value at Six-Month Scan | Value at Twelve-Month Scan |
| --- | --- | --- |
| Age at scan date (years) | 25.6 ± 5.8 | 26.2 ± 5.9 |
| BMI (kg/m<sup>2</sup>) | 23.5 ± 4.0 | 23.4 ± 4.0 |
| Time since ACLR at MRI Scan (months) | 6.9 ± 1.2 | 12.9 ± 0.9 |
| Sex |  |  |
| Male | 10 (40.0%) | 10 (40.0%) |
| Female | 15 (60.0%) | 15 (60.0%) |
| Graft Type |  |  |
| Bone-patellar tendon-bone autograft | 22 (88%) | 22 (88%) |
| Quadriceps tendon autograft | 3 (12%) | 3 (12%) |
| KOOS Score | 75.4 ± 8.6 | 82.0 ± 10.0 |
| Symptoms | 82.7 ± 11.5 | 83.5 ± 13.2 |
| Pain | 85.5 ± 7.8 | 87.6 ± 10.1 |
| Activities of Daily Living | 94.8 ± 5.5 | 97.3 ± 3.7 |
| Sports and Recreation | 61.0 ± 16.2 | 74.4 ± 18.0 |
| Quality of Life | 53.2 ± 15.3 | 66.2 ± 14.9 |
| WOMAC Score | 6.8 ± 5.6 | 5 ± 7.6 |
| Pain | 1.0 ± 1.2 | 1.1 ± 1.7 |
| Stiffness | 1.6 ± 1.2 | 1.0 ± 1.7 |
| Physical Function | 4.2 ± 4.3 | 2.7 ± 4.7 |
| MARS Score | 8.2 ± 5.3 | 9.2 ± 4.5 |

<sup>a</sup>Data are reported as mean ± SD, or n (%). All KOOS scores are on a 0-100 scale, with 0 representing extreme knee problems. All WOMAC scores are on a scale of 0 to 96, with 96 representing extreme knee problems. WOMAC subscore pain is scored from 0-20, stiffness from 0 to 8, and physical function from 0 to 68. All MARS Scores are scaled from 0 to 16 with 16 representing highly physically active individuals. BMI, body mass index. ACLR, anterior cruciate ligament reconstruction. KOOS, Knee Injury and Osteoarthritis Outcome Score. WOMAC, Western Ontario and McMaster Universities Osteoarthritis Index, MARS, Marx Activity Rating (MARS)

### Positive correlations observed between six-month and twelve-month MRI data

The percentage change was mapped on a pixel level from six months to twelve months for each MRI metric (Figure 4). There was a positive correlation between the six-month average shear strain in the femoral central contact area and the twelve-month femoral shear strains (Figure 2, p = 0.02). Furthermore, positive correlations were observed for the axial strains in the tibial cartilage contact areas between the six-month and twelve-month values (Figure 2, p=0.01, p =0.03, respectively). However, no other strain averages displayed significant correlations between their six-month and twelve-month values in any cartilage region. Relaxometry data as a whole exhibited stronger positive correlations between six and twelve month data than strain data. T2 and T2* data showed positive correlations between six-month and twelve-month values in all subregions of the tibial articular cartilage (Figure 3). No significant correlations in T1ρ values between time points were observed in any region other than the femoral central contact region. All other results are displayed in Figure 4.

**Figure 2:**
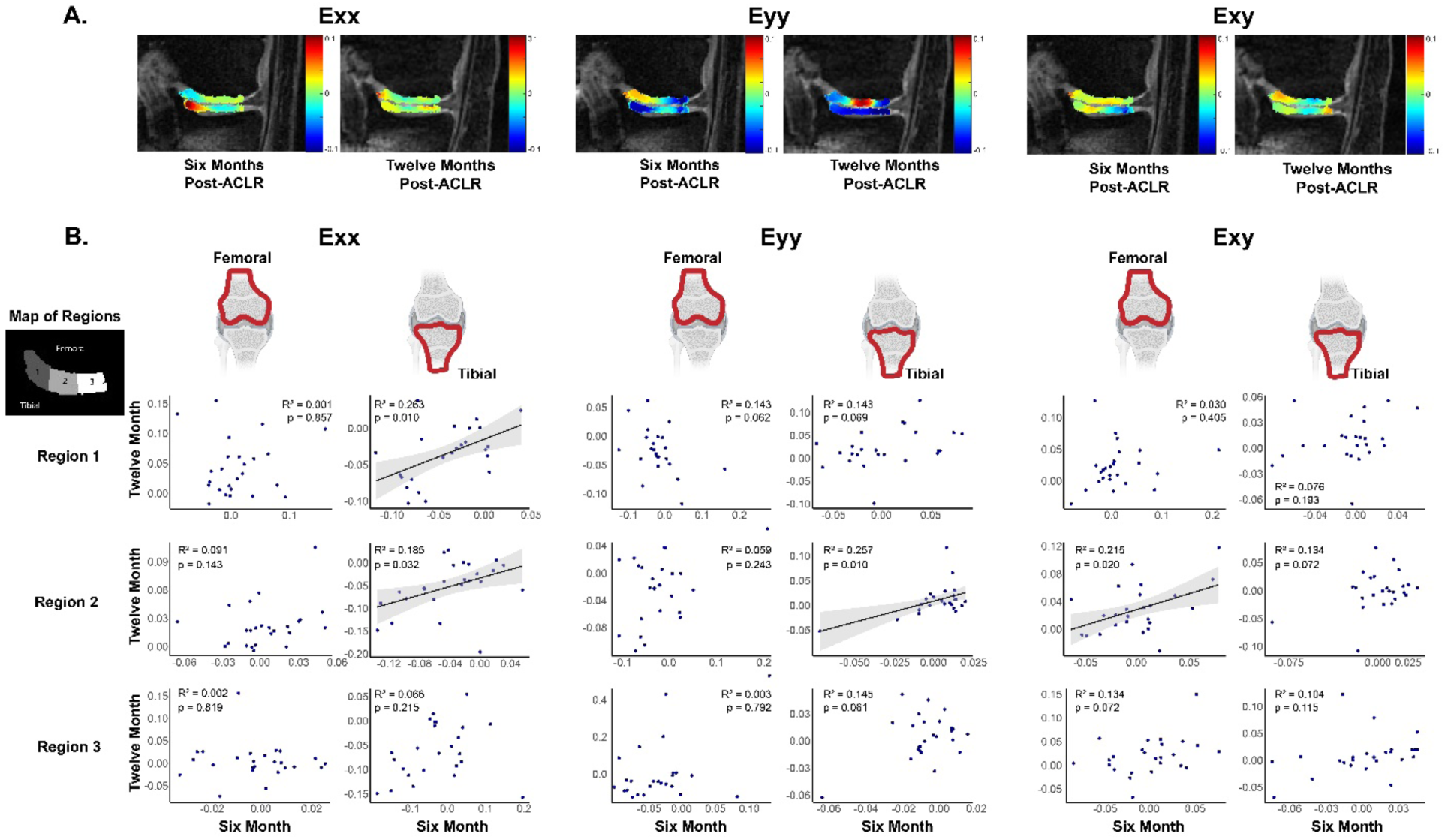
Correlations between six- and twelve-months display variability in time-evolving longitudinal strain patterns. A. Representative images are shown of strain in a single participant at 6 months and 12 months. B. The relationship of average strain values at six- and twelve-months post-ACL reconstruction was evaluated. Most strain measures were not significantly correlated between six-month and twelve-month values. Unadjusted regressions were calculated for the averages in each cartilage subregion. Confidence intervals of 95% are shown in gray. Tibial cartilage experiences significant compression during weight-bearing activities, making axial and transverse strains crucial for assessing its mechanical response post-ACLR. Conversely, femoral cartilage, subjected to shear forces during knee movement, highlights the importance of shear strain in evaluating its structural integrity over time. These findings underscore the complexity of cartilage remodeling dynamics post-ACL surgery.

**Figure 3:**
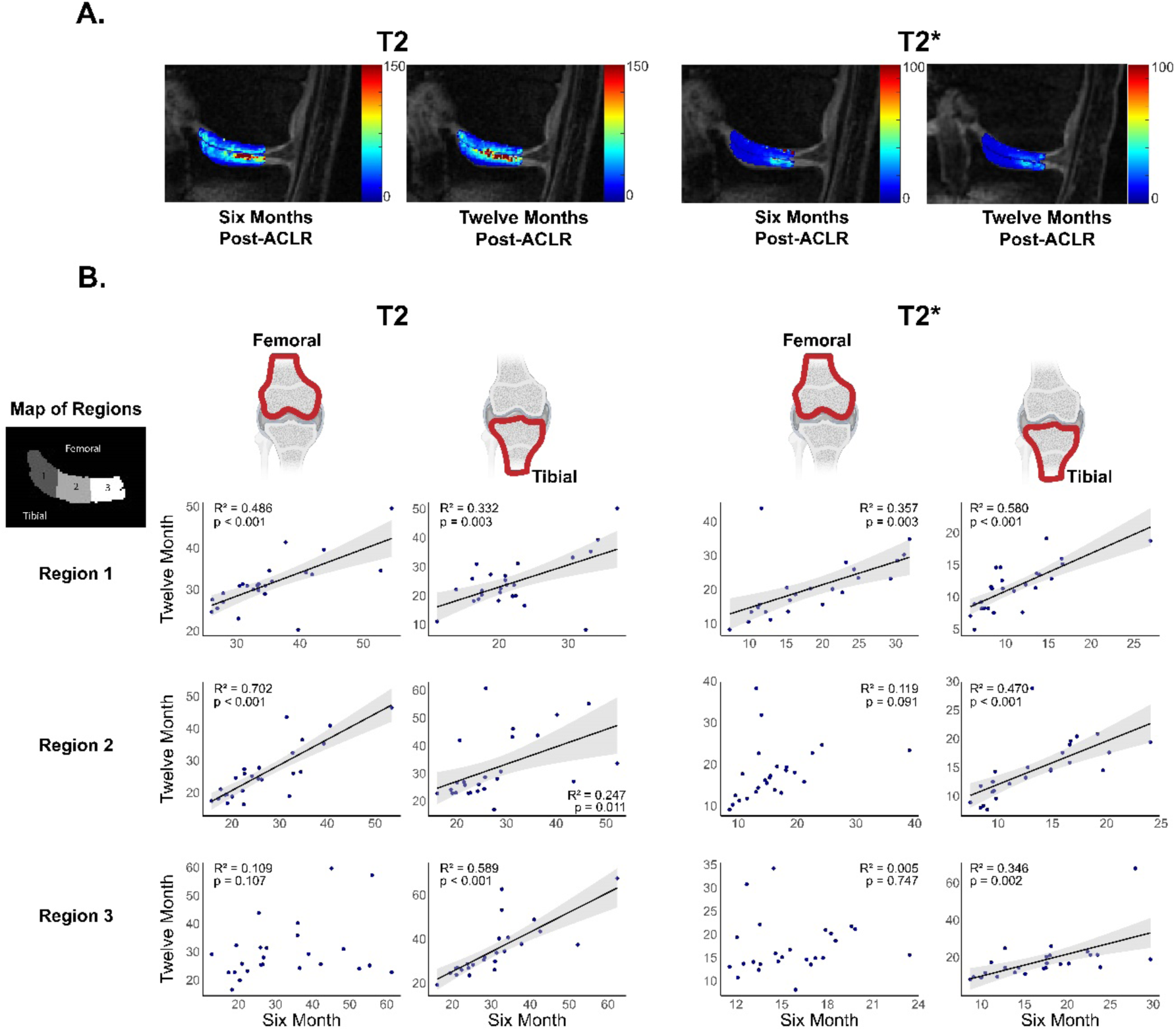
Most relaxometry measures showed significant correlations between their values at six- and twelve-months post-ACLR. Unadjusted regressions were conducted using average values from each cartilage subregion. Gray shading indicates 95% confidence intervals. The inclusion of specific relaxometry measures—T2, T2*, and T1ρ MRI for both femoral and tibial cartilage—reflects their biochemical significance in assessing post-surgical cartilage health. The observed strong correlations suggest that biochemical changes may not yet be detectable over this time interval, underscoring the importance of long-term longitudinal monitoring.

**Figure 4:**
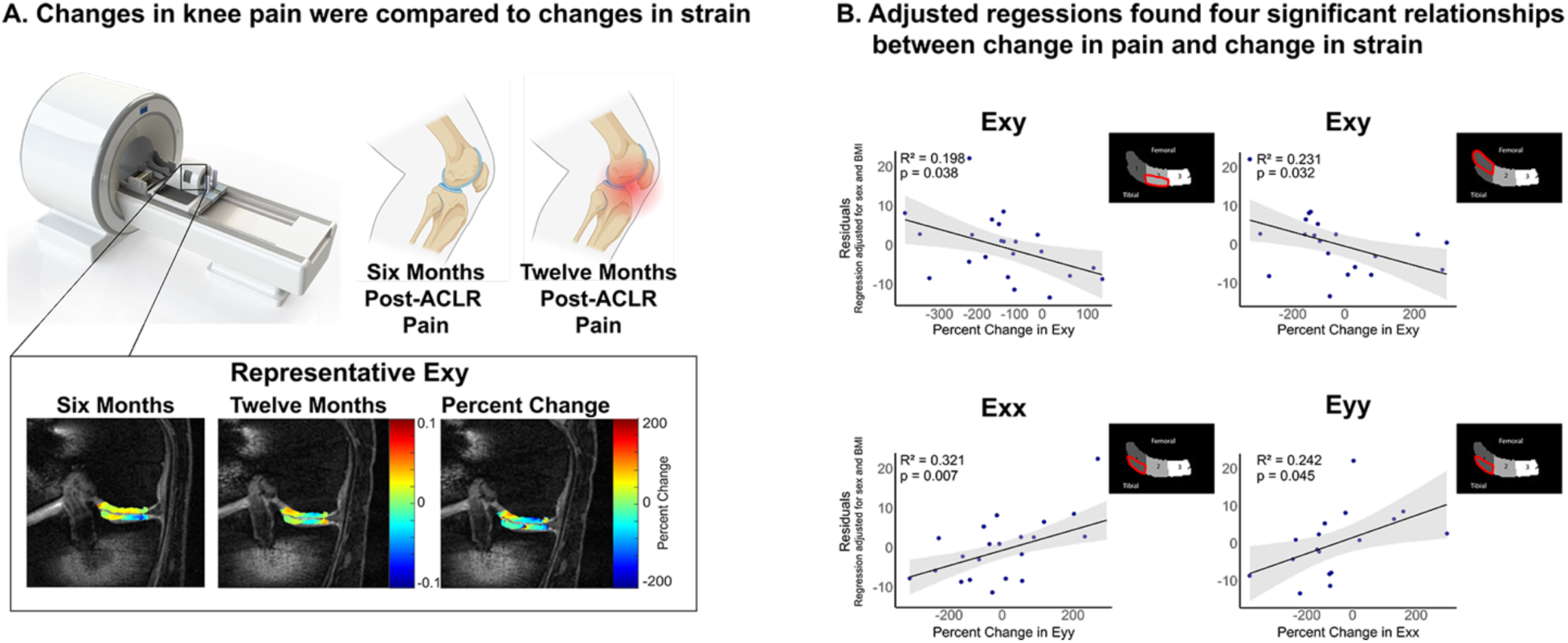
Changes in regional strain patterns were significantly correlated with changes in participant pain, with adjustments made for covariate data such as sex and BMI. Decreased shear strain in the femoral and tibial contact regions correlated with increased pain, whereas increased transverse and axial strain in the tibial contact area also correlated with increased pain. Decreased shear strain suggests altered load distribution and altered cartilage-cartilage contact patterns, potentially exacerbating friction and leading to pain. Conversely, increased transverse and axial strains in the tibial contact area could indicate increased mechanical loads on the cartilage, contributing to pain and cartilage degradation over time. Adjusted regressions are shown, which account for the variability introduced by covariate data such as sex and BMI. Strain is displayed on the horizontal axis and adjusted regression for percent change in pain is displayed on the vertical axis. The pain percent change was determined through the scaling and averaging of the KOOS and WOMAC pain subscores from six to twelve months. No regressions were significant in the outer cartilage-meniscal contact region in either the femoral or tibial cartilage. Confidence intervals of 95% are shown in gray. See supplement Figure S1 for relaxometry data and S2 for nonsignificant strain data.

### MRI changes and pain fluctuations between six and twelve months exhibit variable correlations

The pain percent change was determined through the scaling and averaging of the KOOS and WOMAC pain subscores from six to twelve months. Both unadjusted and adjusted regressions were performed for each cartilage subregion to explore the relationship between participants’ pain change and strain change. The adjusted regressions, presented in Figure 4, controlled for sex and BMI. Four notable correlations were discovered in the analysis of strain data. Specifically, in the inner-most tibial cartilage subregion, it was found that increases in axial and transverse strains between six months and twelve months were associated with increased pain (Figure 4, p = 0.007, p = 0.045, respectively). Conversely, an opposite trend was observed in shear strain. Decreases in shear strain in the inner femoral and central tibial cartilage subregions were linked to increased pain (Figure 4, p = 0.032, p = 0.038, respectively). Notably, no significant correlations were observed in the outer cartilage-meniscal region, as well as with any qMRI data.

### Correlations with the twelve-month knee adduction moment vary by MRI metric

During the loading phase of gait analysis, we calculated the peak KAM for each participant. We conducted both unadjusted and adjusted regressions for each cartilage subregion to investigate the relationship between changes in strain among participants and the 12-month KAM. The adjusted regressions, which are illustrated in Figure 6 for strain data, were controlled for sex and BMI. The analysis of strain data revealed three significant correlations. In the central tibial cartilage region, we observed that increases in axial strain were associated with decreased 12-month KAM (Figure 5, p = 0.029). Conversely, in the same subregion, we found that increases in transverse strain were linked to increased 12-month KAM (Figure 5, p = 0.043). A similar trend was identified in the inner-most femoral cartilage subregion, where decreases in shear strain were associated with increased KAM (Figure 5, p=0.043). Notably, again we did not observe any significant correlations in the outer cartilage-meniscal region or with any qMRI data.

**Figure 5:**
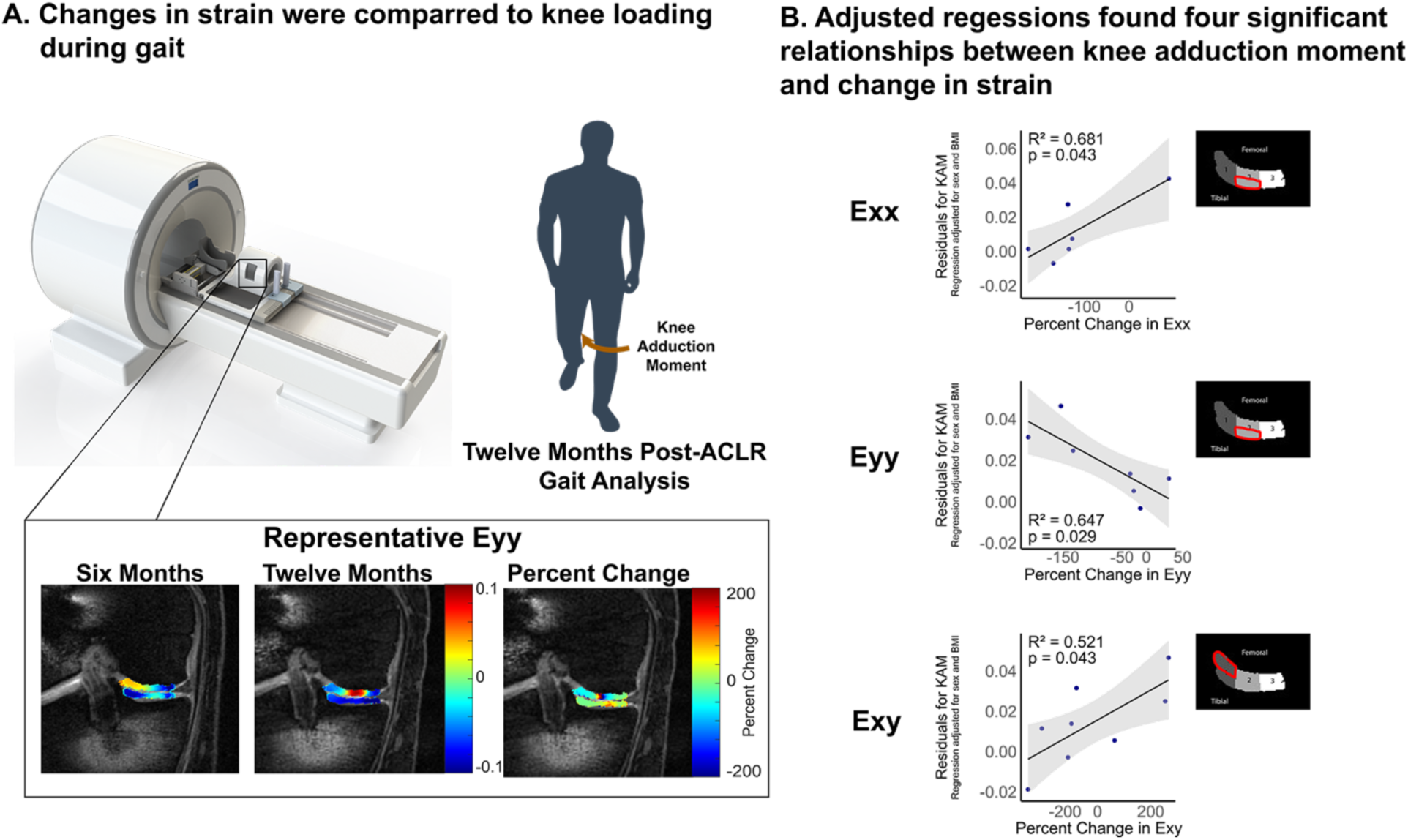
Changes in regional strain patterns were significantly correlated with knee adduction moment. Increases in shear strain in the femoral cartilage contact area were associated with higher knee adduction moments during gait, which was attributed to loss of joint stability and altered load distribution. Alternatively, decreases in axial strain in the tibial contact area of cartilage were correlated with higher knee adduction moments, likely due to realignment of forces through the joint during gait that increased the medial-lateral stress. These findings highlight the complex biomechanical relationships influencing knee joint dynamics and underscore the importance of understanding regional strain patterns using noninvasive imaging measures. Adjusted regressions are shown, which account for the variability introduced by covariate data such as sex and BMI. No regressions were significant in the outer cartilage-meniscal contact region in either the femoral or tibial cartilage. There were no significant regressions found with relaxometry data. Confidence intervals of 95% are shown in gray. All other (nonsignificant) data is shown in Figure S3.

## Discussion

In evaluating the onset of cartilage degeneration, a non-invasive method is critical for monitoring the pre-osteoarthritic cartilage state. Assessing cartilage mechanical and biochemical properties quantitatively could potentially serve as an early indicator of pre-osteoarthritic cartilage changes, which may relate to long-term joint health (24). First, we examined the relationship between medial articular cartilage strains derived from dualMRI as well as qMRI metrics at 6 and 12 months post-ACLR. Second, we assessed how variations in participant pain correlate with changes in MRI metrics over this timeframe. Third, within a subset of participants, we explored the functional implications of strain changes on the peak KAM at 12 months, as quantified by markerless motion capture system. From these objectives. our study yielded three primary outcomes: 1. Increased axial and transverse strains in the tibial cartilage from 6 to 12 months post-ACL reconstruction were significantly correlated with increased pain. 2. Decreases in shear strain in the femoral and tibial cartilage were linked to increases in pain, highlighting the intricate connection between complex biomechanical alterations and cartilage health. 3. Early post-reconstruction regional strain changes were found to be related with knee adduction moment patterns in a representative subgroup population. Our results imply that early post-reconstruction strain patterns are critical for predicting long-term cartilage health.

Our findings align with previous studies that highlight the importance of biomechanical factors in cartilage health (9–14,16,17,25). The use of qMRI and dualMRI techniques provided detailed insights into cartilage strain and relaxometry, offering a more comprehensive understanding of early cartilage changes compared to conventional imaging methods. Additionally, our finding that lower 12-month KAM is associated with decreases in femoral shear strain is supported by other findings(26). Previous work has shown that frontal plane knee kinematics in walking in participants with post-traumatic OA vs non-traumatic OA differs (26). Mechanistically, knee unloading after ACLRmay shift the location and magnitude of force applied to the cartilage during gait, which may disrupt the structural health of cartilage, leading to alterations in regional strain patterns observed via dualMRI (27,28). The association between decreased shear strain in the femoral cartilage contact region and increased pain may suggest that altered load distribution and cartilage-cartilage contact patterns potentially exacerbate friction within the join and lead to pain. Conversely, the relationship between increased transverse and axial strains in the tibial contact area and increased pain could indicate that there are concentrated stresses on the cartilage, contributing to pain and cartilage degradation over time. Additionally, tibial cartilage experiences significant compression during weight-bearing activities, making axial and transverse strains crucial for assessing its mechanical response post-ACLR. Femoral cartilage, subjected to shear forces during knee movement, highlights the importance of shear strain in evaluating its structural integrity over time. Lower KAM impulse at 1 month post-reconstruction is associated with longer T2 relaxation times at 2 years post-surgery, possibly indicating poorer cartilage health (29). However, other studies have shown that increases in joint loading are associated with later presence of OA (30). It is unknown whether initial underloading can lead to progressive overloading or whether it is possible that both lower and higher loads in cartilage may lead to harmful cartilage changes. In our work, the association between increased pain, decreased KAM and specific strain changes indicates a bidirectional relationship where pain may lead to altered gait and further cartilage degradation, or early cartilage changes may increase joint stress and pain, leading to changes in gait.

Modifying the KAM offers a promising strategy for the reduction of pain and alteration of cartilage mechanical properties in individuals with osteoarthritis, in particular following ACLR. Excessive KAM during gait increases the load on the medial compartment of the knee, possibly exacerbating cartilage degradation as well as pain (28). In particular, subject-specific gait modifications that modify the foot progression (e.g., toe-in, toe-out) angle have been shown to reduce peak KAM more than uniformly assigned modifications (31). However, much of the work evaluating gait modifications has been performed in a different patient cohort (e.g., age-related OA), and further work is required to assess the long-term and interventional efficacy of gait modifications in post-traumatic OA, especially in the short-term period one to two years post ACL reconstruction.

Several limitations of this study should be acknowledged. The sample size of 25 participants is relatively small, potentially affecting the generalizability of the results. The same limitation can be applied to the sample size of the gait subset as well. Variability in participant characteristics, such as age, BMI, and rehabilitation adherence, could influence outcomes, although the use of adjusted regressions should allow for some control over introduced variability from covariate data. Additionally, patient-reported outcomes like pain assessments are subjective and may not accurately reflect underlying cartilage health. Future studies with larger, more diverse cohorts and objective measures of pain and function are necessary to validate these findings.

In conclusion, our study underscores the significant impact of altered knee loading on cartilage material properties post-ACLR. The observed regional strain changes and their associations with pain and gait mechanics emphasize the need for early monitoring and intervention to prevent long-term joint degeneration. Advances in qMRI and dualMRI techniques hold promise for detecting early cartilage changes and guiding personalized treatment strategies to improve outcomes for ACLR patients. Future research should focus on larger patient cohorts to further elucidate the mechanisms underlying cartilage degeneration, and KAM as a modifiable intervention through gait retraining that may positively impact the evolution of cartilage health and patient pain.

## Data Availability

All data produced in the present study are available upon reasonable request to the authors.

## Acknowledgments

Financial support was provided by the National Institutes of Health (2 R01 AR063712 and AR063712S1). The funder of the study had no role in study design, data collection, data analysis, data interpretation, or writing of the report.

The authors are thankful for MRI technical support from Teryn S. Wilkes at the Intermountain Neuroimaging Consortium at University of Colorado Boulder.

## List of Abbreviations

ACL: Anterior Cruciate Ligament
ACL: Anterior Cruciate Ligament Reconstruction OA: Osteoarthritis
qMRI: Quantitative Magnetic Resonance Imaging
dualMRI: displacements under applied load Magnetic Resonance Imaging
KAM: Knee adduction moment
KOOS: Knee Injury and Osteoarthritis Outcome Score
WOMAC: Western Ontario and McMaster Universities Arthritis Index

## Strobe Checklist

STROBE Statement—checklist of items that should be included in reports of observational studies

|  | Item No | Recommendation | a(N/A) |
| --- | --- | --- | --- |
| **Title and abstract** |  |  |  |
|  | 1 | (a) Indicate the study's design with a commonly used term in the title or the abstract | yes |
|  |  | (b) Provide in the abstract an informative and balanced summary of what was done and what was found | yes |
| **Introduction** |  |  |  |
| Background/rationale | 2 | Explain the scientific background and rationale for the investigation being reported | yes |
| Objectives | 3 | State specific objectives, including any prespecified hypotheses | yes |
| **Methods** |  |  |  |
| Study design | 4 | Present key elements of study design early in the paper | yes |
| Setting | 5 | Describe the setting, locations, and relevant dates, including periods of recruitment, exposure, follow-up, and data collection | yes |
| Participants | 6 | (a) *Cohort study*—Give the eligibility criteria, and the sources and methods of selection of participants. Describe methods of follow-up<br>*Case-control study*—Give the eligibility criteria, and the sources and methods of case ascertainment and control selection. Give the rationale for the choice of cases and controls<br>*Cross-sectional study*—Give the eligibility criteria, and the sources and methods of selection of participants | yes |
|  |  | (b) *Cohort study*—For matched studies, give matching criteria and number of exposed and unexposed<br>*Case-control study*—For matched studies, give matching criteria and the number of controls per case | yes |
| Variables | 7 | Clearly define all outcomes, exposures, predictors, potential confounders, and effect modifiers. Give diagnostic criteria, if applicable | yes |
| Data sources/<br>measurement | 8* | For each variable of interest, give sources of data and details of methods of assessment (measurement). Describe comparability of assessment methods if there is more than one group | yes |
| Bias | 9 | Describe any efforts to address potential sources of bias | yes |
| Study size | 10 | Explain how the study size was arrived at | yes |
| Quantitative<br>variables | 11 | Explain how quantitative variables were handled in the analyses. If applicable, describe which groupings were chosen and why | yes |
| Statistical methods | 12 | (a) Describe all statistical methods, including those used to control for confounding | yes |
|  |  | (b) Describe any methods used to examine subgroups and interactions | yes |
|  |  | (c) Explain how missing data were addressed | yes |
|  |  | (d) *Cohort study*—If applicable, explain how loss to follow-up was addressed<br>*Case-control study*—If applicable, explain how matching of cases and controls was addressed<br>*Cross-sectional study*—If applicable, describe analytical methods taking account of sampling strategy | yes |
|  |  | (e) Describe any sensitivity analyses | yes |

## Supplemental Materials

**Figure S1:**
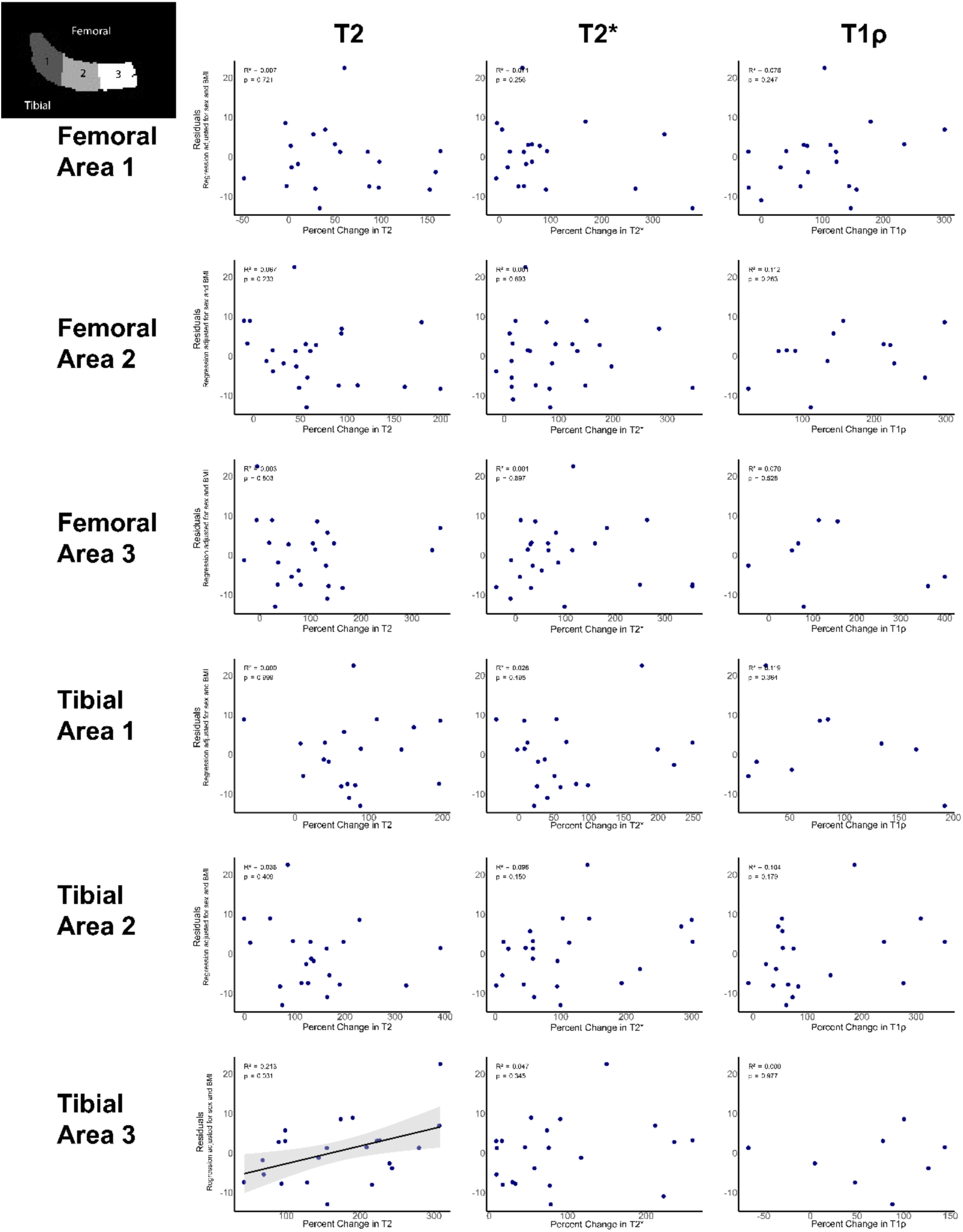
Changes in regional relaxometry values were not significantly correlated with changes in participant pain, with adjustments made for covariate data such as sex and BMI. Only increased T2 values in the tibial cartilage-meniscal contact area displayed a significant positive correlation with participant changes in pain. Increased T2 values could indicated higher water content and changes in collagen organization, signifying early signs of cartilage degeneration. Given that this region plays a crucial role in load transmission during weight-bearing activities, elevated T2 values may indicate that the cartilage is undergoing structural changes in response to increased mechanical stress. Adjusted regressions, adjusted for the variability introduced by covariate data such as sex and BMI are shown. Average relaxometry value is shown on the x-axis and adjusted regression for percent change in pain is shown on the y-axis. The pain percent change was determined through the scaling and averaging of the KOOS and WOMAC pain subscores from six to twelve months. Confidence intervals of 95% are shown in gray.

**Figure S2:**
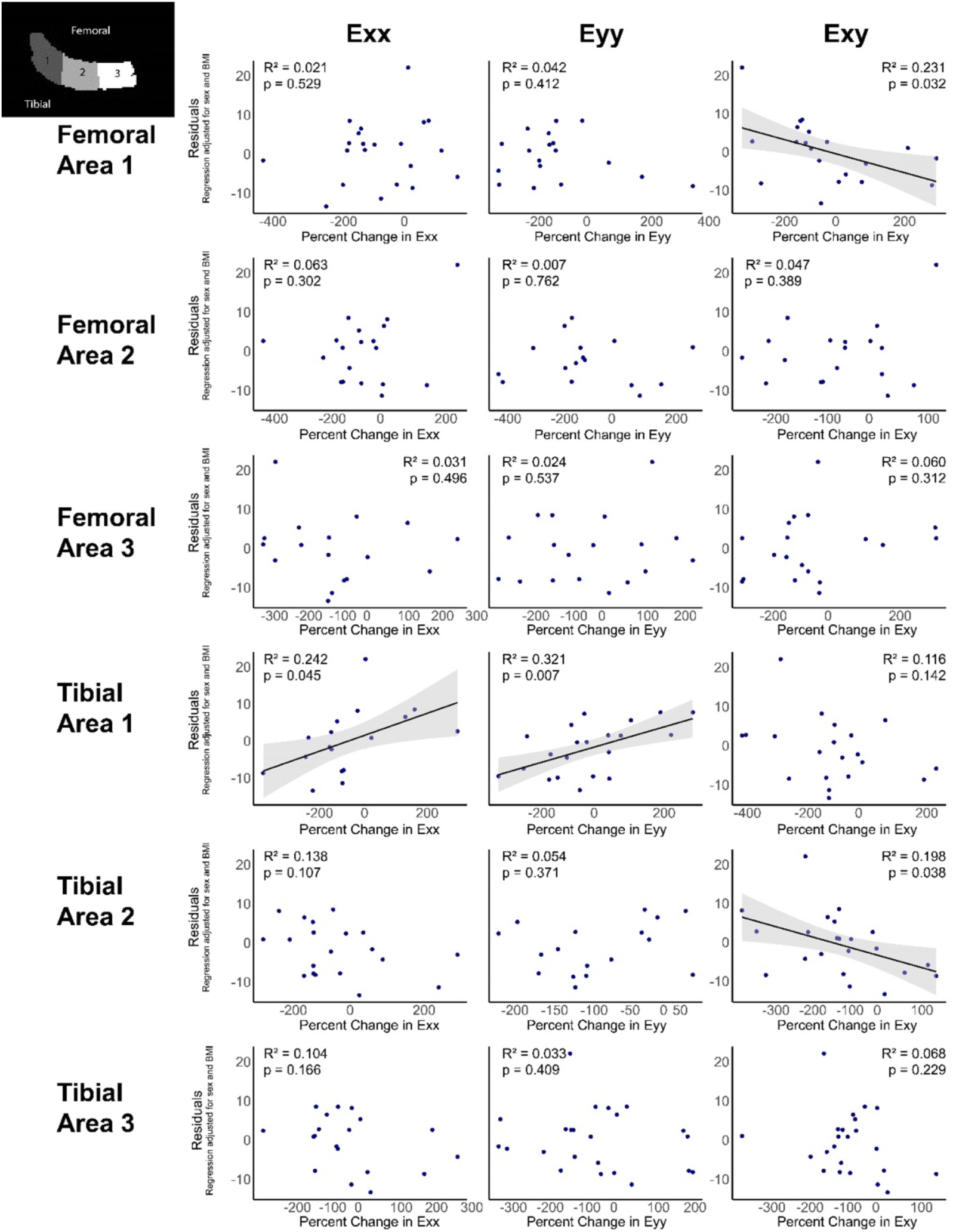
Changes in regional strain patterns were significantly correlated with changes in participant pain, with adjustments made for covariate data such as sex and BMI. All data, both significant and unsignificant is shown here, whereas only the significant relationships are shown in Figure 4.

**Figure S3:**
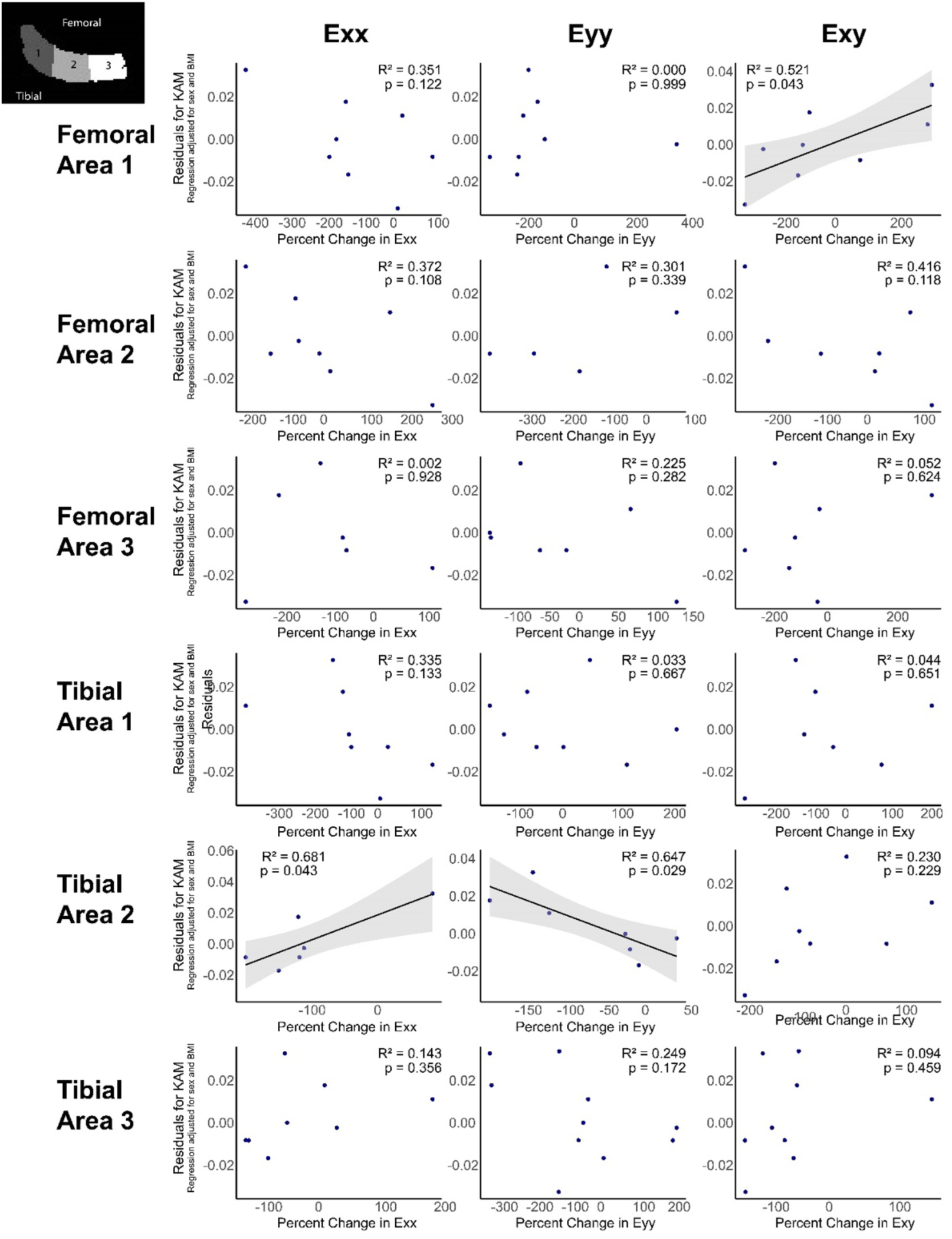
Changes in regional strain patterns were significantly correlated with knee adduction moment. All data, both significant and unsignificant is shown here, whereas only the significant relationships are shown in Figure 5.

